# Proteome-wide Mendelian randomization implicates TIMP2 as a putative causal protein for bone mineral density and fracture risk

**DOI:** 10.64898/2025.11.28.25341226

**Authors:** Chen-Yang Su, Maya Akerman, Masashi Hasebe, Douglas P. Kiel, Satoshi Yoshiji

**Affiliations:** Quantitative Life Sciences, McGill University, Montréal, Québec, Canada; Victor Phillip Dahdaleh Institute of Genomic Medicine, McGill University, Montréal, Québec, Canada; Canada Excellence Research Chair in Genomic Medicine, McGill University, Montréal, Québec, Canada; Department of Biology, McGill University, Montréal, Québec, Canada; Department of Human Genetics, McGill University, Montréal, Québec, Canada; Department of Diabetes, Endocrinology and Nutrition, Kyoto University Graduate School of Medicine, Kyoto, Japan; Hinda and Arthur Marcus Institute for Aging Research, Hebrew SeniorLife, Boston, MA, USA; Department of Medicine, Beth Israel Deaconess Medical Center and Harvard Medical School, Boston, MA, USA; Programs in Metabolism and Medical & Population Genetics, The Broad Institute of MIT and Harvard, Cambridge, MA, USA

**Keywords:** Mendelian randomization, proteomics, circulating proteins, bone mineral density, osteoporosis, fractures, TIMP2, SOST, RSPO3, drug targets

## Abstract

Osteoporosis is a prevalent cause of fractures in older adults and remains a source of morbidity that requires efforts to develop therapeutics. Circulating proteins play a critical role in the pathophysiology of osteoporosis and offer opportunities to identify new causal determinants of bone health. We therefore performed a large-scale proteome-wide Mendelian randomization (MR) analysis to estimate the effects of genetically determined circulating proteins levels on bone mineral density (BMD) and fracture risk. Genetic instruments were derived from *cis*-protein quantitative trait loci (*cis*-pQTLs) for 2,110 plasma proteins across four European ancestry cohorts and applied to genome-wide association studies (GWAS) of heel estimated BMD, femoral neck BMD, lumbar spine BMD, any fracture, and forearm fracture in up to 426,824 individuals of European ancestry. Across proteins and outcomes, 192 protein-skeletal outcome associations showed MR evidence of association, without evidence for heterogeneity or horizontal pleiotropy, and 128 of these further showed strong colocalization with osteoporosis-related loci. We then prioritized proteins that replicated across cohorts, exhibited concordant effect directions, and were likely to be active in circulation, yielding 18 high-confidence causal proteins for BMD and fracture risk. These included established skeletal regulators such as sclerostin (SOST) and R-spondin-3 (RSPO3), which showed opposing effects consistent with their known biology, along with less well-characterized proteins. Higher genetically predicted tissue inhibitor of metalloproteinases 2 (TIMP2) levels was associated with lower BMD and increased forearm fracture risk. Gene-level and variant-level phenome-wide association analyses converged on skeletal traits, and rare predicted damaging or loss-of-function variants in *TIMP2* were associated with higher BMD at the heel, spine and hip. Our findings implicate several circulating proteins as putatively causal factors for osteoporosis and, among them, provide multiple layers of evidence supporting TIMP2 as a genetically supported candidate for further functional and translational evaluation.

**Lay summary:** Osteoporosis is characterized by decreased bone density, and despite available medications, it remains a key risk factor for fractures, requiring continued effort for development of new therapeutics. We used genetic data to estimate the effect of genetically predicted levels of 2,110 blood proteins on strength and fracture risk. We found 18 proteins with effects on bone mineral density and fractures. One protein, tissue inhibitor of metalloproteinases 2 (TIMP2), showed robust evidence linking its higher levels to decreased bone density and increased risk of forearm fracture, highlighting TIMP2 as a promising new treatment target for osteoporosis.

## Introduction

Osteoporosis is a common age-related skeletal disorder characterized by decreased bone mineral density (BMD), leading to an increased risk of fractures. Globally, about 19.7% of adults meet WHO diagnostic criteria for osteoporosis^1^. Each year, an estimated 9 million fragility fractures occur worldwide, including roughly 1.6 million hip fractures, the most severe type of fracture^2^. These consequences are a major cause of chronic pain, disability, and mortality in older adults, and represent a substantial personal and socioeconomic burden. Although current therapies, including antiresorptive and anabolic agents, can reduce fracture risk, for example, bisphosphonates lower vertebral fractures by 50-70%, hip fractures by 40%, and non-vertebral fractures by 20-30%^3^, substantial residual fracture risk remains, and there is still a need to develop new drug treatment that can be safely used for the long term protection against fracture. The most successful new therapeutic strategies often come from targets with a genetic influence on a disease such as osteoporosis.

Circulating proteins are involved in diverse biological processes, and their dysregulation has been linked to many age-related diseases^4–6^. Recent evidence indicates that bone homeostasis, an essential process disrupted in osteoporosis, is also influenced by proteins in bloodstream^7^. Numerous circulating proteins have been associated with BMD and fracture risk in observational studies^8^, but such findings are susceptible to residual confounding and reverse causation, limiting their ability to establish causal relationships. At the same time, circulating proteins represent attractive therapeutic targets because they are accessible to therapeutic modulation, including monoclonal antibodies, small molecules, or protein-based therapies. Their presence in blood also makes them suitable candidates for biomarker development for early disease detection, prognosis, and monitoring of treatment response^9,10^.

Establishing causal relationships in human biology often requires approaches that can overcome the inherent limitations of observational studies. Mendelian randomization (MR) offers such an approach by using genetic variants as instrumental variables to infer causality^11^. Because genetic variants are randomly allocated at conception and remain fixed throughout life, MR is less susceptible to confounding and reverse causation. Recent advances in high-throughput proteomics and genome-wide association studies (GWAS) have enabled proteome-wide MR analyses to evaluate the causal effects of thousands of proteins across a wide range of disease outcomes^5,12–17^, including osteoporosis-related traits^18–22^. However, earlier studies examined only a limited number of cohorts with proteomic data as exposures and limited outcomes^23–26^. Combining multiple cohorts with proteomic data at population-scale sample sizes and broad coverage^4,13,27,28^, together with expanded fracture GWAS datasets^29^, now make it possible to refine and extend earlier findings.

In this study, we applied proteome-wide MR to systematically estimate the causal effects of genetically determined circulating proteins on BMD and fracture risk. By combining the latest large-scale proteomic GWAS with GWAS of osteoporosis-related outcomes, we generated an updated and more comprehensive assessment of protein targets implicated in osteoporosis. Compared with prior proteome-wide MR studies of osteoporosis, our study incorporates several design features intended to improve causal inference and target prioritization. Specifically, we integrate *cis*-pQTL from four large independent proteomic GWAS cohorts measured across SomaScan v4 and Olink Explore plasma proteomics platforms, apply stringent variant-to-gene (V2G) *cis*-pQTL filtering, and evaluate colocalization using three complementary colocalization methods. We further conduct phenome-wide association (PheWAS) analyses, rare variant collapsing analyses, and druggability assessments to identify proteins with consistent genetic support as potential therapeutic targets for osteoporosis.

## Materials and Methods

### MR overview

We performed two-sample proteome-wide MR using circulating proteins as exposures and osteoporosis-related phenotypes as outcomes. For the protein GWAS, we used summary statistics from four of the largest single-cohort plasma-proteome GWAS currently available, each comprising individuals of European ancestry (described below). Summary statistics for osteoporosis outcomes were obtained from five independent BMD and fracture GWAS. All analyses adhered to the STROBE-MR reporting guidelines^11^.

### Proteomic exposure data

Plasma protein GWAS were originally conducted in four independent cohorts of European ancestry: the Atherosclerosis Risk in Communities (ARIC) study (7,213 participants, 4,657 proteins measured with SomaScan v4)^27^, the Fenland study (10,708 participants, 4,775 proteins measured with SomaScan v4)^28^, the deCODE cohort (35,559 participants, 4,907 proteins measured with SomaScan v4)^13^, and the UK Biobank Pharma Proteomics Project (UKB-PPP; 34,557 participants from the European-ancestry discovery cohort, 2,923 proteins measured with Olink Explore 3072)^4^. These datasets were used to define instrumental variables for MR, as described below.

### Osteoporosis outcome data

We assessed five outcomes derived from individuals of European ancestry. The three BMD outcomes were heel quantitative-ultrasound estimated BMD (eBMD; *N* = 426,824) by Morris et al^30^, femoral neck BMD (FN BMD; *N* = 32,735) from Zheng et al^31^, and lumbar spine BMD (LS BMD; *N* = 28,498) from Zheng et al^31^. The two fracture outcomes were any fractures in the UK Biobank (*N* = 426,795; cases = 53,184)^30^ and forearm fractures across eight Northern European biobanks (*N* = 1,021,094; cases = 50,471)^29^.

### Instrument definition and identification

Instruments for the protein exposures were defined according to criteria established in our previous work^16,32^. We use protein quantitative trait loci (pQTLs), which were genetic variants that were significantly associated with plasma protein abundance. To reduce the risk of horizontal pleiotropy, we restricted instruments to *cis*-acting pQTLs (*cis*-pQTLs) within 500 kb of the transcription start site (TSS) of the corresponding protein-coding gene and associated at *P* < 5×10^−8^ because they are more likely to directly influence protein abundance^14,27^. *Cis*-pQTLs mapping to genes in the HLA region (MICA, MICB) and to CD74 were excluded because of the complex linkage disequilibrium (LD) structure and high pleiotropy in this region^33^. LD clumping (*r*^2^ < 0.001, 1 Mb window) was performed separately in each cohort using 50,000 unrelated European participants from the UK Biobank (UKB 50K) as the LD reference. We retained variants with minor allele frequency (MAF) > 0.01.

To further reduce risk of horizontal pleiotropy, we applied an additional “strict V2G” filtering step, as described previously^16,32^. In each cohort, we first retained only variants that were associated in *cis* with a single protein-coding gene (“strict” *cis*-pQTLs). We then evaluated these variants using the Open Targets Genetics variant-to-gene (V2G) scoring system, which integrates multiple lines of evidence to prioritize likely causal genes. A variant was retained only if it had the highest V2G score for its associated protein-coding gene. Variants that passed both criteria formed the final set of strict V2G *cis*-pQTL instruments that were used in the MR analyses. This selection process ensures that each genetic instrument is strongly associated with a single protein-coding gene, thereby minimizing the risk of violating MR assumptions and strengthening the validity of any inferred causal associations with osteoporosis outcomes.

### Two-sample MR

We conducted two-sample proteome-wide MR to estimate the possible causal effects of genetically predicted protein levels on osteoporosis-related outcomes using the TwoSampleMR package (v.0.5.6). For clarity, we hereafter refer to “genetically predicted protein levels” simply by the protein’s name. Potentially causal effects for proteins with a single instrument were estimated using the Wald ratio, and those with multiple instruments were estimated using inverse-variance weighted (IVW). Statistically significant protein-outcome associations were identified using a Benjamini-Hochberg false discovery rate (FDR) threshold of 0.5%. To assess weak instrumental bias, we calculated F-statistics for all instruments and ensured that all exceeded 10.

### Sensitivity analyses

We conducted several sensitivity analyses to assess the robustness of the MR estimates.

1. Alternative MR estimators: For each protein, we re-estimated causal effects using three pleiotropy-robust methods: weighted median, weighted mode, and MR-Egger.
2. Between-instrument heterogeneity: Heterogeneity was assessed using Cochran’s Q statistic (*P* > 0.05) and the corresponding *I*^2^ (< 50%). We classified results as Pass when *I*^2^ < 0.50 or Q-value > 0.05, Fail when *I*^2^ ≥ 0.50 and Q-value ≤ 0.05, and NA for single-instrument models.
3. Directional horizontal pleiotropy: MR relies on the assumption of no horizontal pleiotropy, which requires that the instruments used for MR analyses act on the target outcome exclusively through the exposure of interest. We tested this assumption using the MR-Egger intercept test when three or more instruments were available. Associations with an intercept *P* ≥ 0.05 were considered Pass, whereas those with an intercept *P* < 0.05 were considered Fail, and single-instrument models were classified as NA.

In summary, a protein-outcome association was considered to “pass all sensitivity checks” if it met all of the following criteria: (i) FDR < 0.005; (ii) heterogeneity Pass/NA; and (iii) pleiotropy Pass/NA.

### Colocalization

To assess whether circulating proteins and osteoporosis outcomes share the same underlying causal variant(s), and to mitigate confounding by LD, we employed three colocalization methods: coloc v.6.0.0^34^, PWCoCo v.1.0^31^, and SharePro v.5.0.0^35^. These methods accommodate scenarios with either single or multiple causal variants. A posterior probability for a shared causal variant (PPH4) > 0.8 in any method was considered evidence of colocalization. Protein-outcome associations that passed MR sensitivity analyses and demonstrated colocalization were classified as providing evidence for a causal hypothesis

### Identifying robust and biologically relevant proteins

We applied three additional filters to protein-outcome pairs that passed MR and colocalization:

1. Cross-cohort replication: We retained only associations that passed MR and sensitivity analyses and demonstrated colocalization (PPH4 > 0.8) in at least two of the four cohorts.
2. Direction concordance: Among replicated associations, we required that their effect estimates were directionally concordant across the cohorts (i.e., β-coefficients showed the same sign).
3. Evidence of biological activity in circulation: We performed a literature review to confirm that each remaining protein has documented biological activity in plasma. We operationally defined evidence of biological activity in circulation as published evidence that the protein is secreted and/or detectable in plasma in a biologically active form, with reported systemic effects when altered or modulated (**Supplementary Table 10**).

### Phenome-wide association study of *TIMP2* and lead *cis*-pQTLs

We prioritized TIMP2 for orthogonal follow-up because it showed one of the largest negative MR effects on eBMD and replicated with concordant direction across all four proteomic cohorts with strong colocalization support. To assess whether the TIMP2 signal could plausibly reflect bone-relevant biology, rather than broad pleiotropy, we performed gene- and variant-level phenome-wide association analyses. We queried traits associated with *TIMP2* and the lead *cis*-pQTLs of *TIMP2* (rs11077399, rs9894212, rs931227, rs8066695) using the Association to Function (A2F) Knowledge Portal (https://a2f.hugeamp.org:8000/gene.html?gene=TIMP2), which curates and harmonizes all GWAS summary statistics from the GWAS catalog. A2F uses GWAS summary statistics to calculate gene-level association scores with MAGMA (Multi-marker Analysis of GenoMic Annotation)^36^. For each trait, MAGMA aggregates all common variants within ±50 kb of a gene, adjusts their joint effects for LD using ancestry-matched 1000 Genomes reference panels, and reports a single *P-*value per gene for each ancestry and at the trans-ethnic level. Gene-level associations reaching *P* ≤ 2.5 X 10^−6^ are considered significant.

For variant-level PheWAS, A2F provides trait effect estimates (betas) with respect to the effect allele from each GWAS. When the A2F effect allele differed from the allele used in our harmonized data, we re-oriented the A2F effect estimates to the TIMP2-increasing allele of the corresponding lead *cis*-pQTL to ensure consistency in effect direction across data sources.

### *TIMP2* rare variant collapsing analysis

We obtained *TIMP2* rare variant collapsing results from the AstraZeneca UK Biobank program in individuals of European ancestry using the ptvraredmg model, which aggregates rare protein-truncating and predicted damaging missense variants into a qualifying variant (QV) carrier indicator. We prioritized ptvraredmg a priori because it provides a principled balance between functional specificity and statistical power by enriching for variants most likely to impair *TIMP2* function (PTVs and predicted damaging missense) while maintaining sufficient QV carrier counts for stable estimation, particularly for DXA-derived BMD phenotypes; in contrast, more restrictive PTV-only models may be underpowered due to sparse carriers, whereas more permissive missense models can dilute true effects by including many likely neutral variants.

For each continuous phenotype, we extracted the reported effect size (β in standard deviation [SD] units comparing QV carriers with non-carriers), standard error, 95% confidence interval (CI), two-sided *P-*value, sample size, and counts of QV carriers and non-carriers. Outcomes were prespecified BMD endpoints: heel BMD, lumbar spine (L1-L4), femoral neck (left), and femur total (left). When bilateral measures were available, we prioritized left-sided values to reduce redundancy. Reported associations were summarized without re-estimation of effect sizes and visualized using forest plots showing β and 95% CIs. *P*-values were nominal, and emphasis was placed on the directional concordance of effects across skeletal sites and imaging modalities.

### Druggability assessment

For the prioritized 18 proteins, we calculated the average Z-score of MR estimates across cohorts, capping values at ±10. Heatmaps were generated using pheatmap (R v4.1.2), with row annotations indicating druggability tiers (Tier 1-3) from Finan et al.^37^ and the presence of each protein in the Open Targets and DrugBank databases (Yes/No).

## Results

### Study overview

The study design is shown in **Figure 1**.

**Figure 1.**
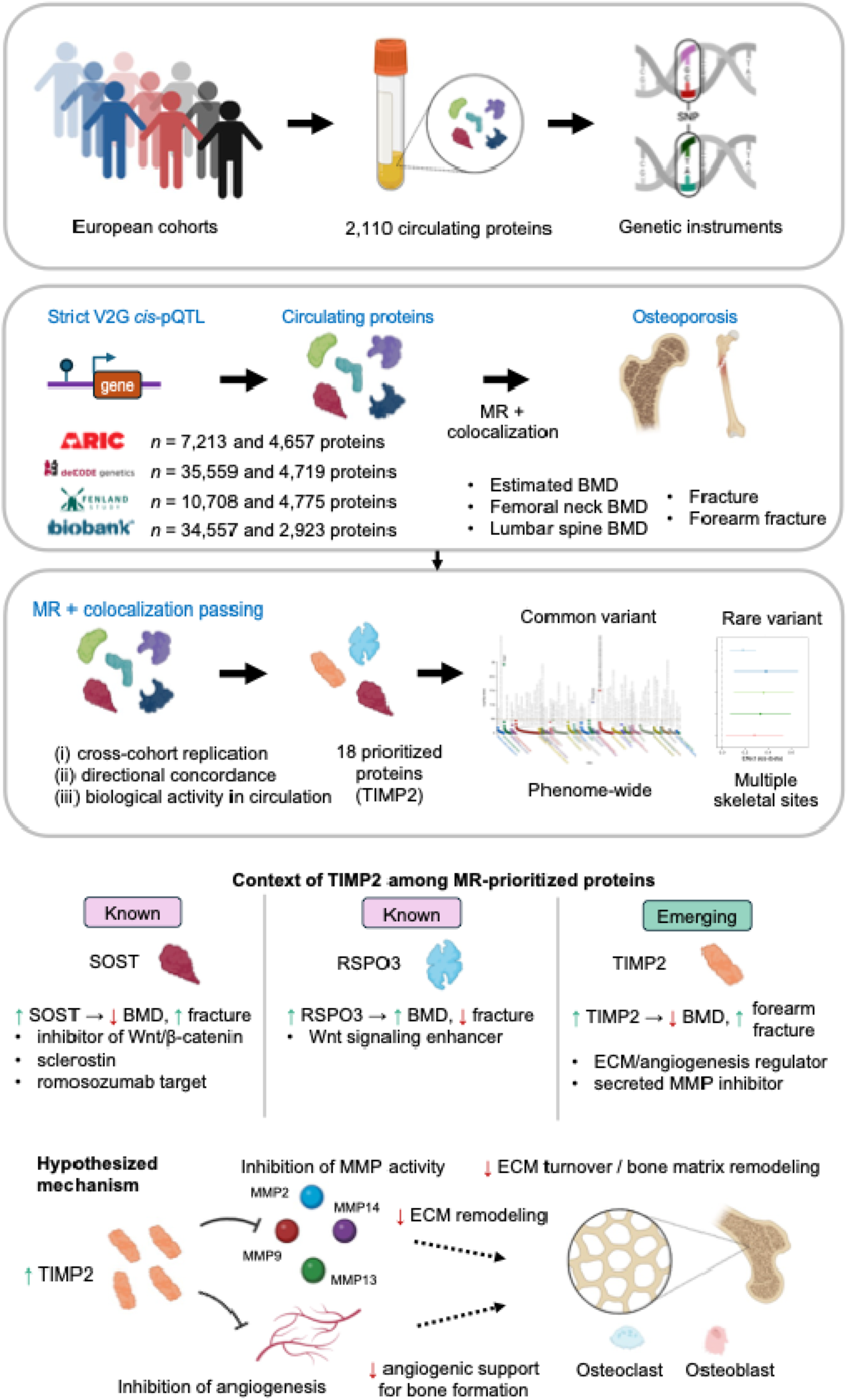
Overview of study design and prioritization of TIMP2 as a therapeutic target for osteoporosis. A) Genetic instrument selection. Proteomic and genetic data from four European-ancestry cohorts (ARIC, deCODE, Fenland, UKB-PPP) were aggregated, and *cis*-pQTLs were curated for 2,110 circulating proteins. Strict variant-to-gene (V2G) filtering was applied to retain genome-wide significant variants that map uniquely to a single protein-coding gene and have the strongest variant-to-gene evidence, yielding 6,224 independent *cis*-pQTL instruments. B) Proteome-wide Mendelian randomization (MR), colocalization, and protein prioritization. Strict V2G *cis*-pQTLs were used as instruments in two-sample MR to estimate the causal effects of circulating proteins on five osteoporosis-related outcomes: heel estimated bone mineral density (BMD), femoral neck BMD, lumbar spine BMD, any fracture, and forearm fracture. Associations passing multiple-testing correction and MR sensitivity analyses were taken forward to colocalization (coloc, PWCoCo, SharePro) to assess whether protein and outcome signals shared a causal variant. High-confidence proteins were then prioritized using three criteria: (i) cross-cohort replication, (ii) directional concordance of effect estimates, and (iii) evidence of biological activity in circulation, resulting in 18 MR- and colocalization-supported proteins, including TIMP2 (tissue inhibitor of metalloproteinases 2). *TIMP2* was further characterized using common-variant phenome-wide association analyses and rare-variant collapsing analyses of bone traits across multiple skeletal sites. C) Context of TIMP2 among MR-prioritized proteins. Among the 18 prioritized proteins, SOST and RSPO3 acted as internal biological benchmarks with well-established opposing effects on bone (SOST: higher protein levels associated with lower BMD and higher fracture risk; RSPO3: higher protein levels associated with higher BMD and lower fracture risk). TIMP2 emerged as a novel candidate, with higher genetically predicted circulating TIMP2 associated with lower BMD and increased forearm fracture risk, whereas rare damaging or loss-of-function TIMP2 variants were associated with higher BMD. D) Proposed mechanism for TIMP2 in bone remodelling. TIMP2 is a secreted inhibitor of matrix metalloproteinases (including MMP14, MMP2, and MMP13) that regulate extracellular matrix (ECM) turnover and angiogenesis. Elevated TIMP2 is hypothesized to reduce MMP activity, leading to impaired ECM remodelling, altered osteoclast–osteoblast coupling, and reduced angiogenic support for bone formation, thereby lowering BMD. Conversely, genetic reductions in TIMP2 activity are associated with increased BMD.

### Genetic instrument selection

The genetic instrument selection process is shown in **Supplementary Figure 1**. We curated *cis*-pQTLs from four European ancestry cohorts^4,13,27,28^ (**Supplementary Table 1**). In total, 6,224 independent *cis*-pQTLs instrumenting 2,110 circulating proteins (1,106 in ARIC, 1,247 in deCODE, 1,198 in Fenland, 1,294 in UKB-PPP) remained after LD clumping and our strict V2G criteria. Briefly, strict V2G *cis*-pQTLs are genome-wide significant variants associated with a single protein-coding gene and showing the strongest variant-to-gene support according to the Open Targets Genetics V2G score. There was no evidence of weak instrument bias (median F-statistics = 255, 298, 233, 448 across ARIC, deCODE, Fenland, and UKB-PPP cohorts, respectively; range 30–13,425). (**Supplementary Table 2**).

### Two-sample proteome-wide MR

We estimated the causal effects of circulating proteins on osteoporosis-related traits using two-sample MR. Strict V2G *cis*-pQTLs served as instrumental variables, and the five outcomes were eBMD, FN BMD, LS BMD, any fracture, and forearm fracture (**Supplementary Table 1**). For each cohort, harmonized protein-outcome summary statistics used for MR are provided in **Supplementary Tables 3-6**, respectively. Using a Benjamini Hochberg-corrected FDR threshold of 0.5% in each outcome for each cohort, we identified 192 unique protein-outcome associations (180 for eBMD; 2 for FN BMD; 2 for LS BMD; 2 for fracture; 6 for forearm fracture) that met MR and sensitivity analysis criteria, including heterogeneity and directional horizontal pleiotropy (**Supplementary Table 7**).

### Colocalization and prioritization of high-confidence proteins

To reduce the possibility that MR results were biased by LD, we conducted colocalization using three methods (coloc, PWCoCo and SharePro). A posterior probability for a shared causal variant (PPH4 > 0.8) in any method was obtained for 128 protein-outcome pairs (117 for eBMD; 2 for FN BMD; 1 for LS BMD; 2 for fracture; 6 for forearm fracture), representing 67% of MR-significant associations (**Supplementary Table 8**).

To identify robust and biologically relevant proteins, we applied three criteria: (i) cross-cohort replication, (ii) directional concordance, and (iii) documented biological activity in circulation. Forty-six protein-outcome associations involving 42 proteins replicated across cohorts with concordant effect directions (**Supplementary Table 9**). Of these, 22 associations spanning 18 proteins had evidence of biological activity in circulation: ANGPTL4, C5, CTSS, EGF, FRZB, GDF15, ITIH3, KNG1, OLFML3, PLAU, PLTP, RSPO3, SERPINF2, SERPING1, SMOC2, SOST, TIMP2, and TGFBI (**Supplementary Table 10**). The corresponding 22 associations are provided in **Supplementary Table 11**.

Two well-established skeletal regulators, SOST (sclerostin) and RSPO3 (R-spondin-3), served as internal validation markers. As expected from their known biological roles^38–40^, the two proteins showed strong but opposing associations with BMD and fracture risk. SOST, which suppresses osteoblast activity, was negatively associated with eBMD (β_UKB-PPP_ = −0.51, *P*_UKB-PPP_ = 6.1 × 10^−59^; **Figure 2**) and FN BMD (β_Fenland_ = −0.59, *P*_Fenland_ = 9.4 × 10^−8^; **Figure 3a**), and was positively associated with both overall fracture risk (β_Fenland_ = 0.096, *P*_Fenland_ = 2.7 × 10^−12^; **Figure 3b**) and forearm fracture risk (β_Fenland_= 0.10, *P*_Fenland_ = 6.9 × 10^−25^; **Figure 3b**). RSPO3, a stimulator of osteoblast differentiation via Wnt signaling, showed positive associations with eBMD (β_Fenland_ = 0.21, *P*_Fenland_ < 2.3 × 10^−308^; **Figure 2**) and was negatively associated with both overall fracture risk (β_Fenland_ = −0.18, *P*_Fenland_ = 8.3 × 10^−21^; **Figure 3b**) and forearm fracture risk (β_ARIC_ = −0.22, *P*_ARIC_ = 9.1 × 10^−51^; **Figure 3b**). For each protein, we report the association with the lowest *P*-value observed across cohorts for simplicity.

**Figure 2.**
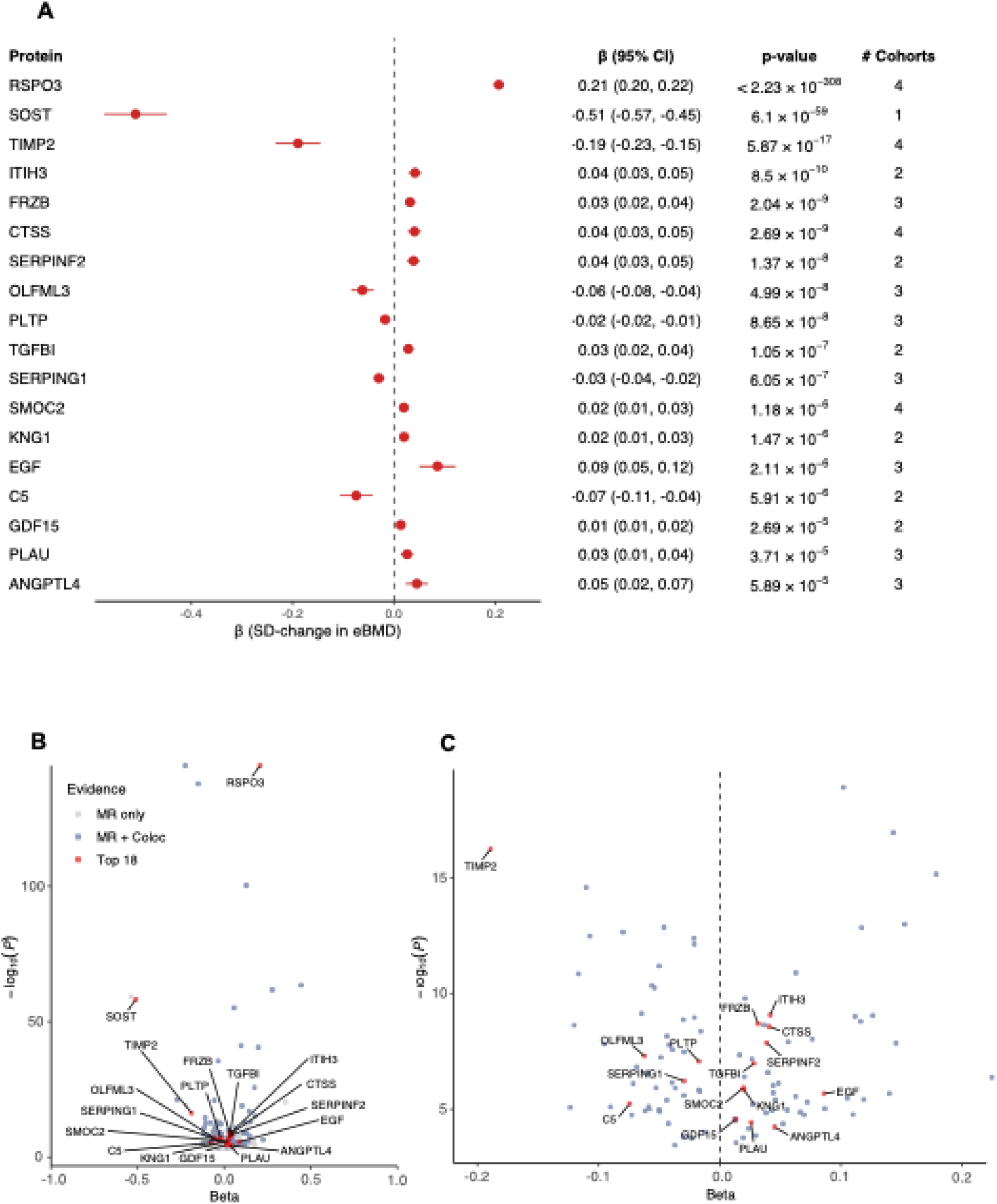
Proteome-wide Mendelian randomization of putatively causal circulating proteins with evidence of biological activity in circulation with heel estimated BMD (eBMD). (A) Forest plot of the 18 MR- and colocalization-prioritized proteins with evidence of biological activity in circulation. For each protein, the point denotes the MR effect estimate (β, SD change in eBMD per 1 SD higher genetically predicted protein level) from the cohort with the smallest *P-*value, and the horizontal bar shows the 95% confidence interval (CI). The accompanying table reports β (95% CI), *P*-value, and the number of cohorts with proteomic data in which the protein-eBMD association passed MR and colocalization criteria. Positive β values indicate higher eBMD and negative β values indicate lower eBMD; SOST and TIMP2 show the largest negative effects, whereas RSPO3 shows a strong positive effect. The number of cohorts indicates the number of cohorts with proteomics (ARIC, deCODE, Fenland, UKB-PPP) in which the protein-outcome association met the prespecified MR sensitivity and colocalization criteria. (B) Volcano plot of all proteins showing MR associations with eBMD at FDR ≤ 0.5% across the four cohorts with proteomic data. The *x*-axis shows the MR effect estimate (β) and the y-axis shows −log_10_(*P*). Points are colored by level of evidence: proteins passing MR only (grey), proteins passing both MR and colocalization (blue), and the top 18 prioritized proteins with evidence of biological activity in circulation shown in panel A (red), with selected proteins labelled. (C) Zoomed view of the volcano plot in panel B, highlighting the position of the 18 prioritized proteins with evidence of biological activity in circulation, including TIMP2, SOST, and RSPO3. For visualization, the displayed estimate corresponds to one representative cohort-specific MR estimate with the smallest *P*-value.

**Figure 3.**
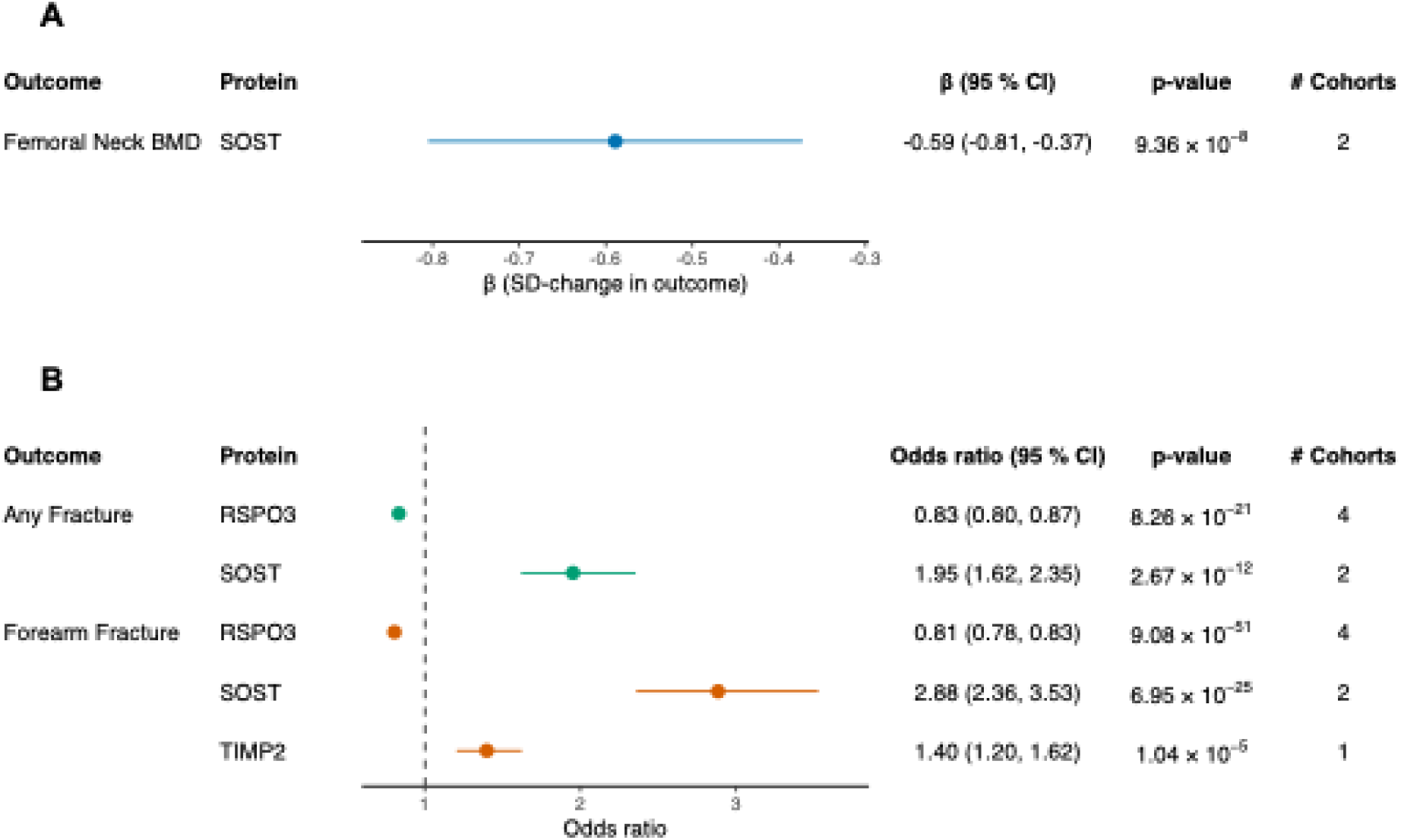
Mendelian randomization (MR) estimates of SOST, RSPO3, and TIMP2 on femoral neck bone mineral density (BMD) and fracture risk. (A) Forest plot showing the association between genetically predicted circulating SOST and femoral neck BMD (SD units). The point represents the MR effect estimate (β) and the horizontal bar indicates the 95% confidence interval (CI) from the cohort with the smallest *P-*value (Fenland); the table indicates β (95% CI), *P-*value, and the number of proteomic cohorts in which the SOST-femoral neck BMD association passed MR and colocalization criteria. (B) Forest plot showing MR-derived odds ratios (ORs) for any fracture and forearm fracture per 1 SD higher genetically predicted protein level for RSPO3, SOST, and TIMP2. Points represent ORs and horizontal bars show 95% CIs, with the vertical dashed line denoting the null (OR = 1). For each protein-outcome pair, the estimate from the cohort with the smallest *P-*value is displayed, together with its OR, 95% CI, *P-*value, and the number of cohorts in which the association replicated. RSPO3 shows protective associations with fracture risk, whereas SOST and TIMP2 show increased fracture risk, consistent with their directions of effect on BMD. All displayed associations passed MR sensitivity analyses and colocalized (PPH4 > 0.8); protein-outcome associations that did not pass these criteria are not shown. The number of cohorts indicates the number of cohorts with proteomics (ARIC, deCODE, Fenland, UKB-PPP) in which the protein-outcome association met the prespecified MR sensitivity and colocalization criteria. For visualization, the displayed estimate corresponds to one representative cohort-specific MR estimate with the smallest *P*-value. Colors denote different outcomes.

Among the less characterized proteins, TIMP2 (tissue inhibitor of metalloproteinases 2) emerged as a compelling target. TIMP2 is a secreted inhibitor of matrix metalloproteinases (MMPs) that regulates extracellular matrix turnover and independently suppresses endothelial-cell proliferation, making it an important modulator of tissue remodelling in both health and disease. At an FDR threshold of 0.5%, a one SD increase in genetically predicted circulating TIMP2 was associated with lower eBMD in all four cohorts with proteomic data (β_ARIC_ = −0.085, β_deCODE_ = −0.19, β_Fenland_ = −0.15, β_UKB-PPP_ = −0.14, *P*_ARIC_ = 1.08 × 10^−7^, *P*_deCODE_ = 5.9 × 10^−17^, *P*_Fenland_ = 1.4 × 10^−9^, *P*_UKB-PPP_ = 1.1 × 10^−12^; **Figure 2**), with consistent direction and broadly similar effect sizes across cohorts. These associations were supported by strong colocalization evidence (PPH4_ARIC_ = 1.00, PPH4_deCODE_ = 1.00, PPH4_Fenland_ = 0.913, PPH4_UKB-PPP_ = 1.00). Additionally, a one SD increase in genetically predicted TIMP2 levels was associated with a higher risk of forearm fracture in the UKB-PPP cohort (β_UKB-PPP_ = 0.34, *P*_UKB-PPP_ = 1.04 × 10^−5^, PPH4_UKB-PPP_ = 0.90; **Figure 3b**), consistent with the direction of the eBMD findings. The lead *cis*-pQTL for TIMP2 differed across cohorts, consistent with differences in proteomic measurement platforms (Somascan vs. Olink) and underlying population genetic architecture: rs11077399 in ARIC, rs9894212 in deCODE, rs931227 in Fenland, and rs8066695 in UKB-PPP.

### Phenome-wide association analyses for *TIMP2*

To obtain orthogonal evidence, we examined *TIMP2* gene- and variant-level associations in the A2F Knowledge Portal, which reports MAGMA-based gene-level scores. Common variants at *TIMP2* showed significant gene-level associations with eBMD (*P* = 2.96 × 10^−18^), height (*P* = 6.34 × 10^−29^), and phosphate levels (*P* = 1.76 × 10^−13^), all relevant skeletal traits (**Figure 4** and **Supplementary Table 12**).

**Figure 4.**
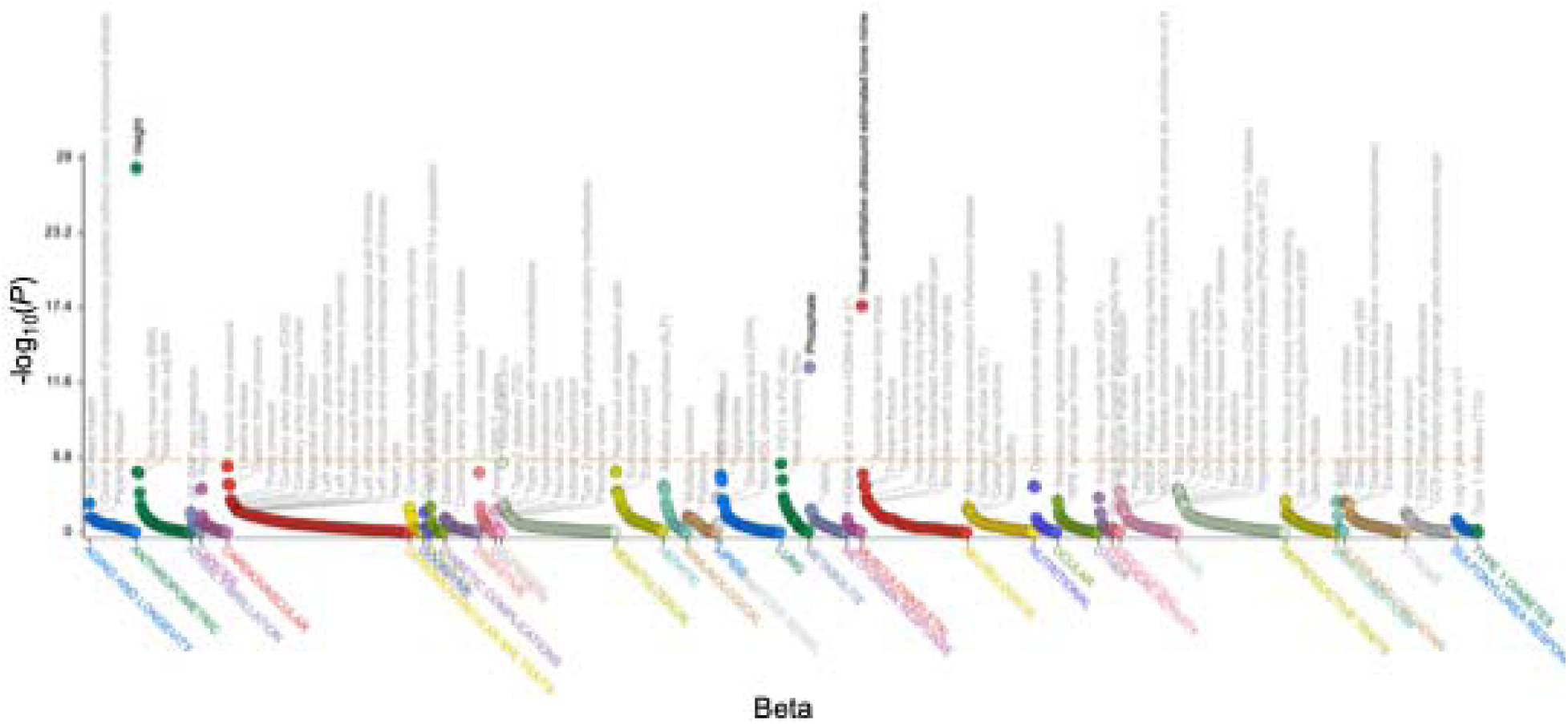
Phenome-wide association results for *TIMP2* based on common-variant gene-based associations from mixed-ancestry gene-level analyses in the Association-to-Function (A2F) Knowledge Portal. Each point represents the MAGMA gene-level association between *TIMP2* and a complex trait, plotted by the effect estimate (β; x-axis) and −log_10_(*P*) (y-axis). Traits are grouped by phenotype category and are distinguished by color. The horizontal orange dashed line indicates the significance threshold used by A2F (*P* ≤ 2.5 × 10^−6^).

We next examined variant-level associations for each cohort’s lead *cis*-pQTL (ARIC: rs11077399; deCODE: rs9894212; Fenland: rs931227; UKB-PPP: rs8066695). Across cohorts, alleles that increased circulating TIMP2 were consistently associated with lower eBMD, whereas TIMP2-decreasing alleles were associated with higher eBMD, mirroring the MR directionality (**Supplementary Figure 2-5**). When effects were aligned to the TIMP2-increasing allele for rs11077399, rs931227, and rs8066695, these variants showed inverse associations with eBMD (rs11077399: β = −0.012, *P* = 4.4 × 10^−9^; rs931227: β = −0.015, *P* = 9.5 × 10^−12^; rs8066695: β = −0.014, *P* = 7.6 × 10^−13^) and height (rs11077399: β = −0.010, *P* = 3.9 × 10^−25^; rs931227: β = −0.011, *P* = 2.1 × 10^−15^; rs8066695: β = −0.010, *P* = 3.6 × 10^−32^). The effect allele of rs931227 was positively associated with body mass index (β = 0.011, *P* = 2.0 × 10^−9^), while the effect allele of rs8066695 was negatively associated with phosphate levels (β = −0.016, *P* = 1.2 × 10^−14^). In contrast, the TIMP2-decreasing allele at rs9894212 was associated with higher eBMD (β = 0.016, *P* = 1.65 × 10^−12^) and greater height (β = 0.006, *P* = 7.75 × 10^−10^), consistent with our MR results. Full per-variant PheWAS results are provided in **Supplementary Table 13-16**.

Taken together, the gene- and variant-level PheWAS signals were enriched for skeletal and skeletal-adjacent traits; however, we observed additional associations, notably BMI and phosphate, for some cohort-specific lead *cis*-pQTLs. These observations motivate cautious interpretation and follow-up analyses to disentangle skeletal-specific from systemic pathways.

### *TIMP2* rare variant collapsing analysis on BMD measures in the UK Biobank

In the AstraZeneca UK Biobank gene-level collapsing analysis of European ancestry participants, carriers of rare, predicted damaging or loss-of-function (LoF) variants in *TIMP2* had higher BMD across multiple skeletal sites (**Supplementary Table 17** and **Figure 5**). Heel BMD was higher in carriers (β = 0.18, 95% CI: 0.065 to 0.29; *P* = 2.1 × 10^−3^; *N* = 262,318; QV = 273). Lumbar spine (L1-L4) was also higher (β = 0.38, 95% CI: 0.10 to 0.66; *P* = 7.9 × 10^−3^; N = 43,010; QV = 42). At the left hip, femoral neck (β = 0.36, 95% CI: 0.098 to 0.62; *P* = 7.0 × 10^−3^; N = 45,212; QV = 47), and femur total (β = 0.28, 95% CI: 0.029 to 0.53; *P* = 0.029; N = 45,145; QV = 47) were all higher in carriers than in non-carriers. Together, these results indicate a consistent but modest protective effect of rare, predicted damaging *TIMP2* variants on BMD across measurement modalities.

**Figure 5.**
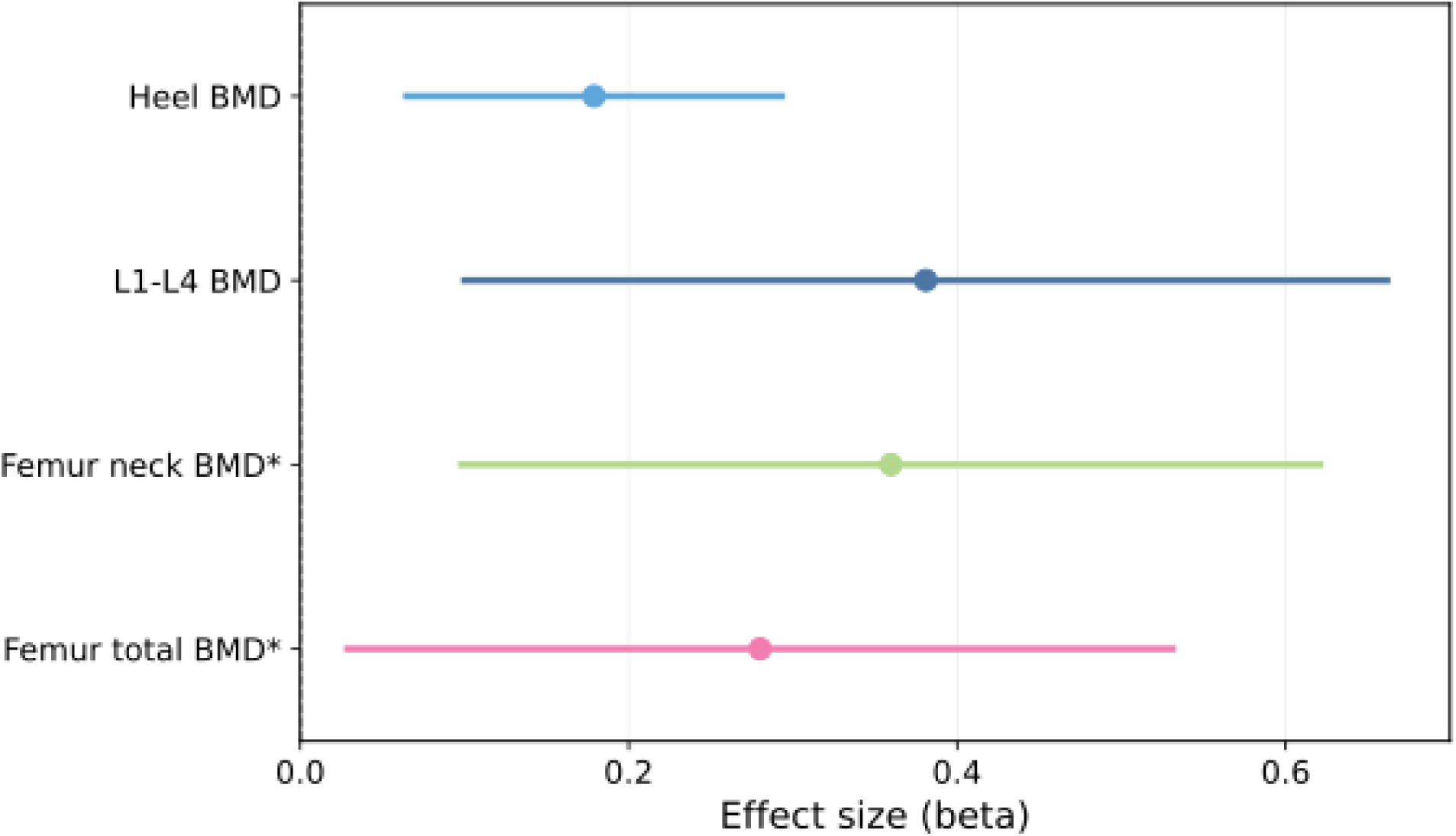
Associations between collapsing analysis of rare damaging or loss-of-function (LoF) *TIMP2* variants and bone mineral density (BMD) outcomes. Forest plots of the associations between carriage of qualifying rare variants in *TIMP2* (ptvraredmg collapsing model) and BMD outcomes in European-ancestry individuals. Outcomes are four prespecified endpoints: heel BMD, lumbar spine L1-L4, femoral neck, and femur total. Points indicate regression coefficients (β) and horizontal lines show 95% confidence intervals, with positive β values indicating higher BMD among carriers of a qualifying variant compared with non-carriers. *Left-sided skeletal endpoints are shown.

### Druggability assessment of MR-prioritized proteins

We assessed druggability of the 18 MR- and colocalization-prioritized proteins using three complementary resources: the druggable genome classification (Tiers 1–3), DrugBank, and the Open Targets Platform. The druggable genome framework stratifies proteins by druggability potential, DrugBank catalogs approved and investigational drugs and their targets, and Open Targets links targets to diseases and clinical development stages.

All 18 proteins mapped to the druggable genome, with three proteins (SOST, C5, CTSS) in Tier 1 and one (PLAU) in Tier 2 (**Figure 6**). Half of the 18 proteins had approved or investigational drugs in DrugBank, including established drugged targets such as SOST (target of the anti-sclerostin antibody romosozumab), C5 (targeted by complement inhibitors such as eculizumab and ravulizumab), and CTSS (with multiple small-molecule inhibitors in clinical or preclinical development), and four were listed as targets in the Open Targets Platform (**Figure 6**). Notably, TIMP2 was a Tier 3 secreted protein that did not appear in DrugBank or Open Targets, indicating a lack of prior therapeutic targeting but suggesting therapeutic potential given its extracellular activity and strong genetic evidence for a causal role in bone.

**Figure 6.**
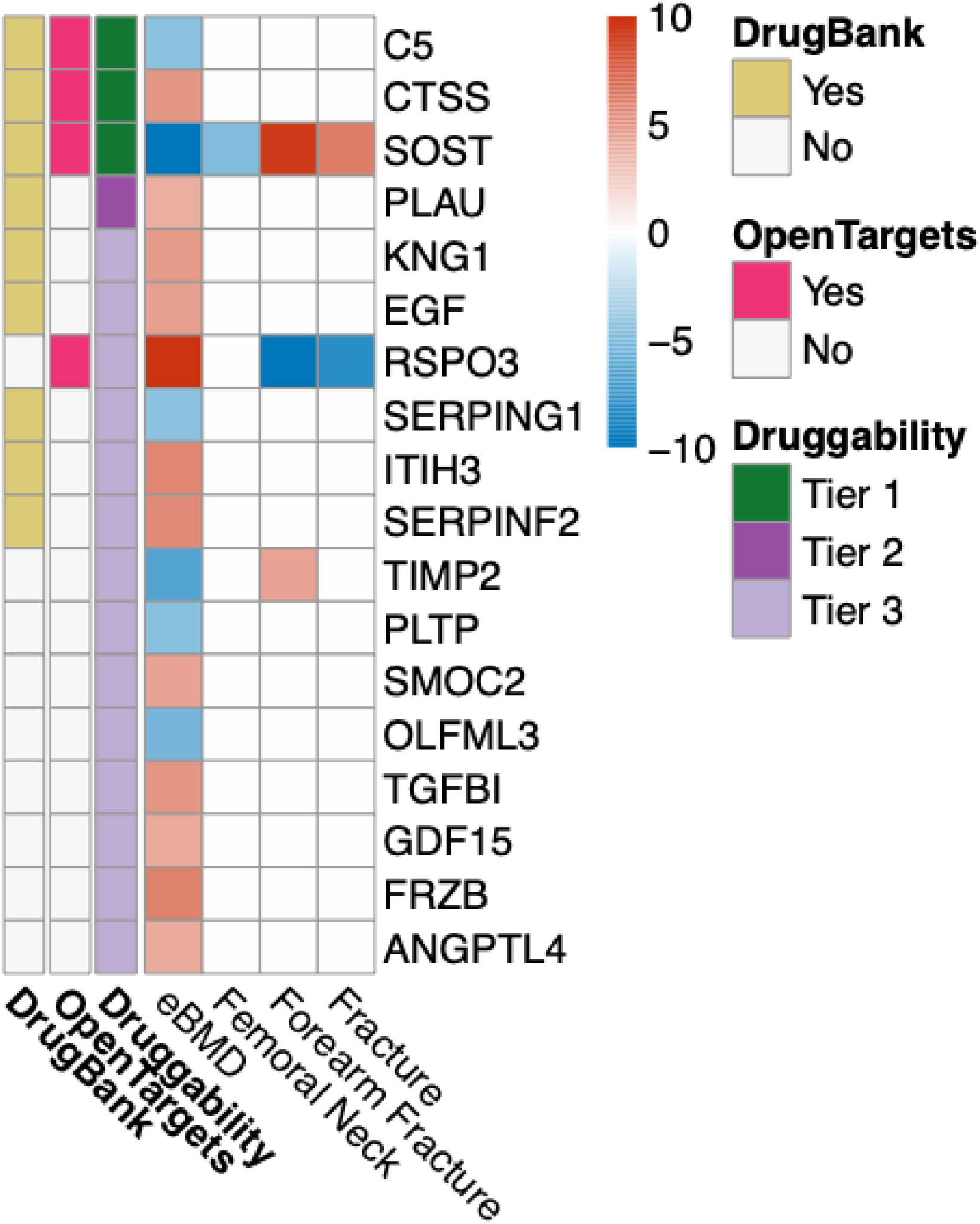
Mendelian randomization (MR) associations between circulating proteins and bone phenotypes with druggability annotations. Heatmap showing Z-scores for the association between genetically predicted circulating levels of 18 MR-prioritized proteins and four outcomes: estimated bone mineral density (eBMD), femoral neck BMD, forearm fracture, and any fracture. Each cell represents the mean MR Z-score for a given protein-outcome pair across European-ancestry cohorts, truncated at ±10. Red and blue denote positive and negative Z-scores, respectively. White cells indicate that the tested protein-outcome pair did not meet the prespecified MR sensitivity analyses and colocalization criteria. Proteins (rows) are ordered by druggability tier from Finan et al. (Tier 1– 3)^37^ and, within tiers, by the amount of external drug evidence. Row annotations indicate the Finan druggability tier and whether the corresponding target is listed in Open Targets or DrugBank (Yes/No).

## Discussion

In this study, we performed large-scale proteome-wide MR analyses and identified 18 circulating proteins with putative causal effects on BMD and fracture risk. These analyses recapitulated two established regulators of skeletal homeostasis, SOST and RSPO3, and also highlighted less characterized proteins, most notably TIMP2, as a compelling candidate. PheWAS further showed that genetically predicted TIMP2 levels are enriched for skeletal traits, and exhibit minimal associations with non-skeletal phenotypes, supporting a predominantly skeletal-trait-related role for TIMP2.

Our MR and gene-based collapsing analyses support an inhibitory role of TIMP2 in skeletal mineralization. Higher genetically predicted circulating TIMP2, reflecting reduced MMP activity, was associated with lower BMD, consistent with recent findings^41^, whereas rare LoF or predicted damaging TIMP2 variants were associated with higher BMD. Across heel, lumbar spine, and left hip, carriers of such variants exhibited higher BMD measures. These collapsing results were directionally concordant with the MR findings for eBMD and collectively support a model in which reduced TIMP2 activity is associated with greater BMD at clinically relevant skeletal sites. The consistency across heel, spine, and hip, argues against a site-specific artifact and instead suggests a generalized skeletal effect of TIMP2 inhibition.

TIMP2 is a secreted inhibitor of MMPs, a family of zinc-dependent peptidases that mediate extracellular matrix (ECM) degradation^42^. MMP activity, particularly that of MMP2, MMP9, MMP13, and MMP14, modulates several key steps of bone remodelling^43^. In mice, targeted deletion of these MMPs that regulate bone remodelling leads to reduced bone mineralization and severe skeletal abnormalities^44–47^, demonstrating that adequate MMP activity is required for normal skeletal development. Consistent with these experimental data, in humans biallelic loss-of-function variants in MMP2 cause hereditary skeletal disorders such as multicentric osteolysis with nodulosis and arthropathy (MONA), Torg, and Winchester syndromes^48–52^. TIMP2 is expressed by osteoblast-lineage cells^53^ and regulates MMP-mediated ECM turnover and osteoclast activity in a dose- and context-dependent manner. At higher concentrations, TIMP2 acts as a broad MMP inhibitor with strong affinity for MMP-2, thereby suppressing bone resorption^54^. At lower concentrations, however, TIMP2 participates in the MMP14-dependent activation of pro-MMP2^55^, which can enhance resorptive activity^56^. Since skeletal homeostasis requires a balanced MMP2-MMP14-TIMP2 axis, chronically elevated TIMP2 may uncouple bone formation and resorption by excessively inhibiting matrix turnover. There are currently no approved osteoporosis therapies that directly target extracellular matrix remodeling pathways.

TIMP2 is also unique among TIMP family members in its ability to directly inhibit endothelial cell proliferation independently of MMP inhibition^57,58^, suggesting that elevated TIMP2 could additionally impair angiogenic support for bone formation. Although dedicated TIMP2-knockout studies remain limited, TIMP2-deficient mice develop accelerated osteoarthritis driven by increased angiogenesis^59^. Consistent with this, the Mouse Genome Informatics database reports that *Timp2*-mutant mice exhibit reduced bone mineral content. TIMP2 expression also declines with age in osteoblasts and osteocytes in mouse long bones^60^, suggesting that TIMP2 may contribute to the maintenance of skeletal integrity during aging. However, this observation is correlative and does not establish a causal role.

At first glance, some prior observations may appear to tension with our genetic findings, but these lines of evidence are not necessarily contradictory. Our MR analyses estimate the effect of lifelong genetically influenced differences in circulating TIMP2, whereas animal knockout models often reflect developmental and tissue-wide perturbation, which can produce qualitatively different phenotypes. In addition, TIMP2 biology is highly context- and dose-dependent within the MMP2-MMP14-TIMP2 axis, and circulating effects may differ from local effects within the bone microenvironment. Finally, some animal readouts such as bone mineral content can be influenced by body size or linear growth, which is notable given the height associations observed in our PheWAS. Accordingly, we interpret our results as genetic evidence consistent with a detrimental effect of higher circulating TIMP2 on BMD, while acknowledging that tissue-specific and developmental roles of TIMP2 may differ.

Our study provides genetic evidence consistent with a causal effect of circulating TIMP2 on BMD and fracture risk. Taken together with existing experimental and observational data, these findings support a model in which increased systemic TIMP2 reduces BMD through systemic over-inhibition of MMP-mediated bone remodelling and potentially diminished angiogenic support for bone formation. Further mechanistic work is warranted to delineate how TIMP2 regulates skeletal homeostasis.

In addition to TIMP2, our study re-identified known skeletal regulators SOST and RSPO3, which served as internal positive controls. SOST encodes sclerostin, an osteocyte-derived secreted glycoprotein that negatively regulates bone formation by inhibiting Wnt/β-catenin signalling and thereby suppressing osteoblast differentiation and activity^39,40^. Sclerostin is the target of the approved anabolic therapy romosozumab used to treat severe post-menopausal osteoporosis. Sclerostin’s known role is consistent with our findings, which show its negative association with BMD and increased fracture risk. Conversely, RSPO3 encodes R-spondin-3, a secreted ligand that stimulates osteoblast activity and promotes bone formation by acting as an enhancer of Wnt signalling^38^. As expected, it showed a positive association with BMD and decreased fracture risk.

Among the other replicated proteins, CTSS (cathepsin S) and SMOC2 (SPARC related modular calcium binding 2) showed consistent positive associations with BMD across all four cohorts. CTSS is a secreted cysteine protease that has previously been implicated in bone remodeling^61^. Although CTSS and C5 (complement component 5) scored highly in druggability assessments, their broad immunological roles and extensive pleiotropy make them less attractive therapeutic targets for osteoporosis^61,62^. SMOC2 is a matricellular protein whose deficiency in mice has been linked to impaired periodontal bone healing and age-related skeletal deterioration, consistent with its positive association with BMD^63^.

Several additional proteins not previously implicated in bone biology, including ITIH3, PLTP, SERPINF2, SERPING1, and OLFML3, also showed consistent causal associations with BMD in our analyses. These findings raise the possibility that they represent novel, uncharacterized regulators of skeletal homeostasis and warrant further functional investigation.

This study has several strengths. First, we integrated data from more than 100,000 individuals across four proteomics cohorts measured on complementary platforms (SomaScan and Olink), enabling both cross-cohort and cross-platform validation. TIMP2 showed a consistent association with reduced eBMD in all four cohorts, providing strong support for a causal role in BMD regulation. We recognize that SomaScan and Olink can differ in epitope and aptamer binding, detection limits, and assay-specific biases, which may lead to imperfect concordance across platforms; however, by requiring directional replication across cohorts and platforms, we adopted a conservative strategy that reduces the risk of platform-specific false-positive findings, albeit at the cost of potentially increasing false negatives (i.e., missing true associations that are not reliably measurable on both platforms). Second, we also extended previous proteome-wide MR studies of osteoporosis by evaluating a broader set of clinically relevant outcomes, including multiple site-specific BMD measures (eBMD, FN BMD, LS BMD) and both general and forearm fracture. Third, we implemented stringent “strict V2G” filtering to select genetic instruments, ensuring that each *cis*-pQTL was strongly and uniquely mapped to a single protein-coding gene and thereby reducing the risk of horizontal pleiotropy. Finally, we applied three state-of-the-art colocalization methods that account for both single and multiple causal variants, mitigating confounding by LD and increasing confidence in the MR results.

This study also has some limitations. First, our MR and colocalization analyses were restricted to individuals of European ancestry. Osteoporosis-related GWAS are still predominantly derived from individuals of European ancestry, and available non-European datasets remain relatively small and underpowered, limiting their suitability for MR. Second, we could not perform sex-stratified analyses because sex-specific proteomic and outcome summary statistics were unavailable, even though sex differences in osteoporosis risk are well-established^1^. Third, fracture GWAS remain less powered than BMD GWAS, which may have contributed to the smaller number of fracture associations detected. Lastly, our work is limited to genetic analyses and does not include functional validation. Follow-up experimental studies and, ultimately, clinical trials will be necessary to confirm the therapeutic potential of targeting TIMP2.

In conclusion, by integrating proteome-wide MR with sensitivity analyses, colocalization, PheWAS, and rare-variant collapsing analyses, we identified 18 proteins with evidence for causal effects on BMD and osteoporosis risk, including the established regulators SOST and RSPO3 and the emerging candidate TIMP2. Our findings highlight TIMP2 as a genetically supported, candidate target for skeletal disease that warrants further functional and translational evaluation.

## Acknowledgements

C.-Y.S. is supported by a CIHR Canada Graduate Scholarship Doctoral Award (Funding Reference Number: 187673), an FRQS doctoral training scholarship, and a Lady Davis Institute/TD-Bank Scholarship. M.H. is supported by a Graduate Scholarship for Degree-Seeking Study Abroad from the Japan Student Services Organization (JASSO) under the special priority framework for doctoral studies at top-tier global universities in STEM fields, and by the 6th Toshizo Watanabe International Scholarship from The Watanabe Foundation. The funders had no role in the study design, data collection and analysis, decision to publish, or preparation of the manuscript. The Yoshiji Lab is supported by the Canada Research Chairs Program (CRC-2025-00097), the Canadian Institutes of Health Research (183596), the DNA to RNA (D2R) Foundational Program, Japan Society for the Promotion of Science, and McGill University.

## Author contributions

**Chen-Yang Su:** Conceptualization; data curation; formal analysis; methodology; software; validation; visualization; writing – original draft; writing – review and editing.

**Maya Akerman**: Formal analysis; visualization; writing – original draft; writing – review and editing.

**Masashi Hasebe:** Writing – review and editing.

**Douglas P. Kiel:** Conceptualization; writing – review and editing.

**Satoshi Yoshiji:** Conceptualization; writing – review and editing; supervision.

## Data Availability

MR results can be accessed in the Supplementary Tables. All MR analyses adhered to STROBE-MR guidelines^11^ (**Supplementary Note 1**).

## Supplementary Figures

**Supplementary Figure 1.**
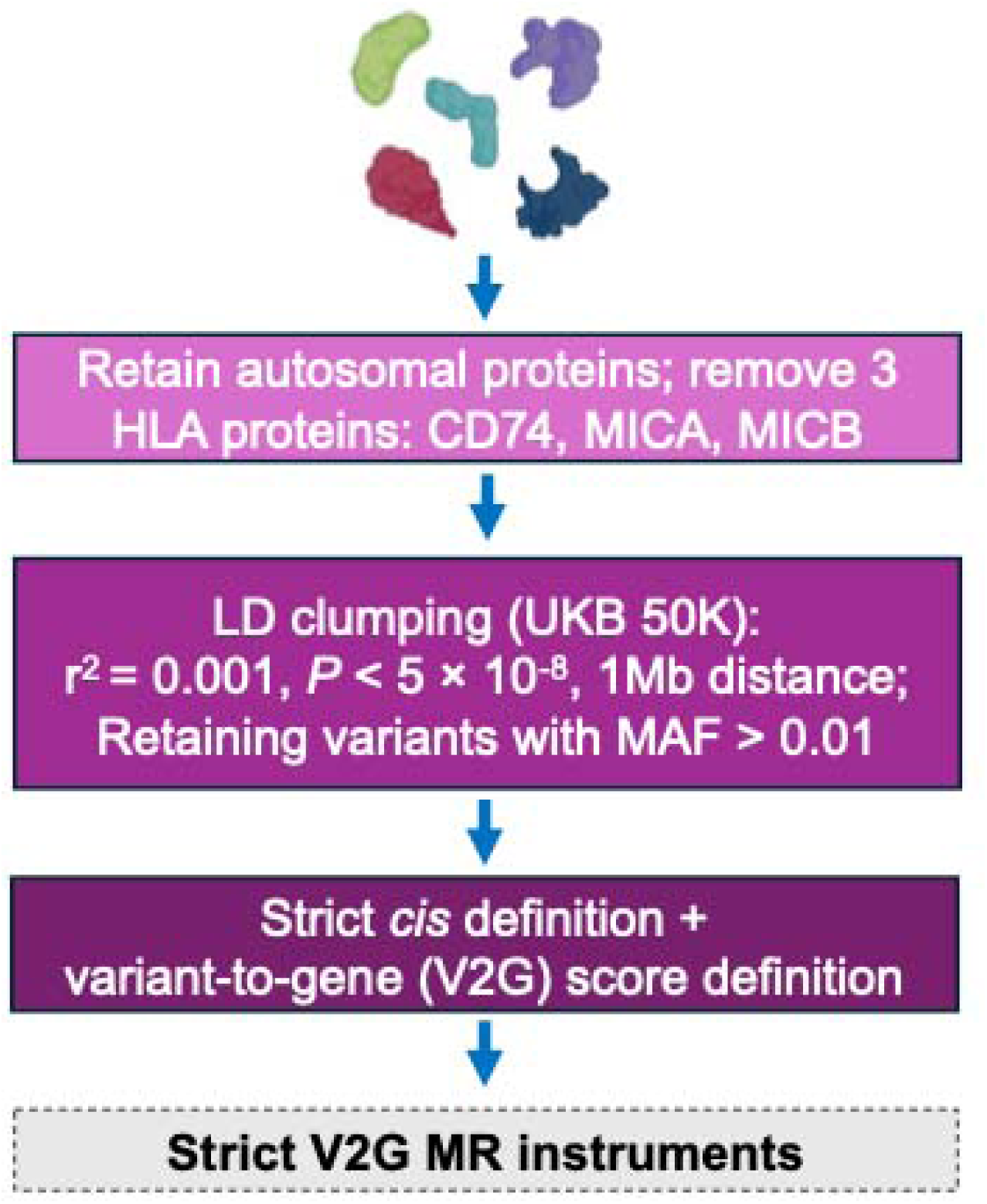
Definition and selection of strict variant-to-gene (V2G) *cis*-pQTL instruments for proteome-wide Mendelian randomization (MR). Flowchart summarizing the instrument identification pipeline. For each proteomics cohort, we first retained autosomal proteins and excluded pQTLs mapping to CD74, MICA, and MICB in the HLA region due to complex linkage disequilibrium (LD) and pleiotropy. We then restricted to genome-wide significant *cis*-pQTLs (within 500 kb of the protein-coding gene, *P* < 5 × 10^−8^) and performed LD clumping (*r*^2^ < 0.001, 1 Mb window) using 50,000 unrelated European individuals from UK Biobank as reference, retaining variants with minor allele frequency > 0.01. Finally, we applied a “strict V2G” filter, keeping only variants that (i) were *cis*-acting on a single protein-coding gene and (ii) had the highest Open Targets Genetics V2G score for that gene. Variants passing all steps comprised the strict V2G *cis*-pQTL used in downstream MR analyses.

**Supplementary Figure 2.**
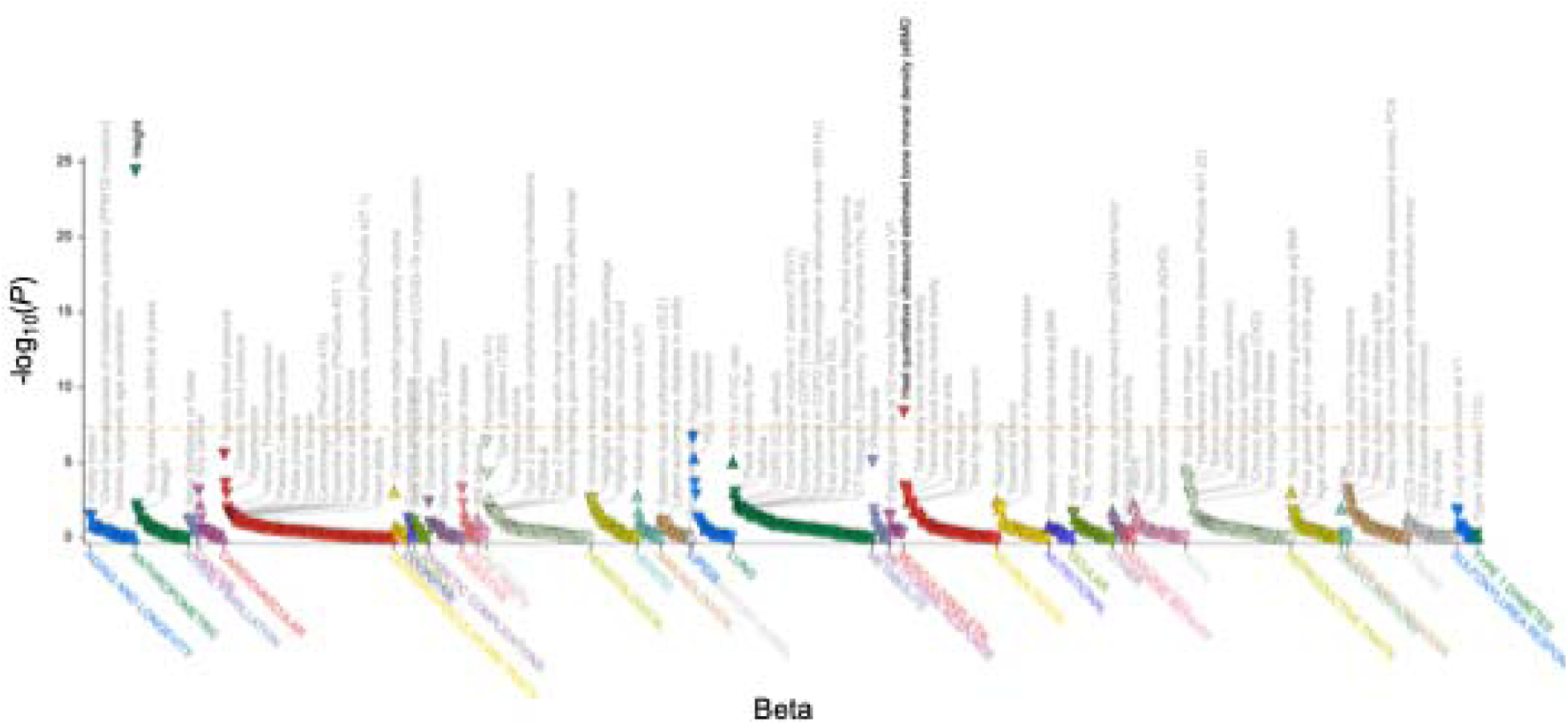
Phenome-wide association results for *TIMP2* lead *cis*-pQTL 17:76864175 A>G (rs11077399) in the ARIC cohort. The plot displays −log_10_(*P*) for the association between the ARIC lead *cis*-pQTL 17:76864175 A>G (rs11077399) and a broad range of phenotypes in the Association-to-Function (A2F) portal. Triangles denote variant-phenotype associations, grouped by phenotype category on the x-axis and distinguished by color. The y-axis shows −log_10_(*P*) for the association test. The horizontal orange dashed line indicates the phenome-wide significance threshold used in A2F; points above the line represent associations significant after multiple-testing correction.

**Supplementary Figure 3.**
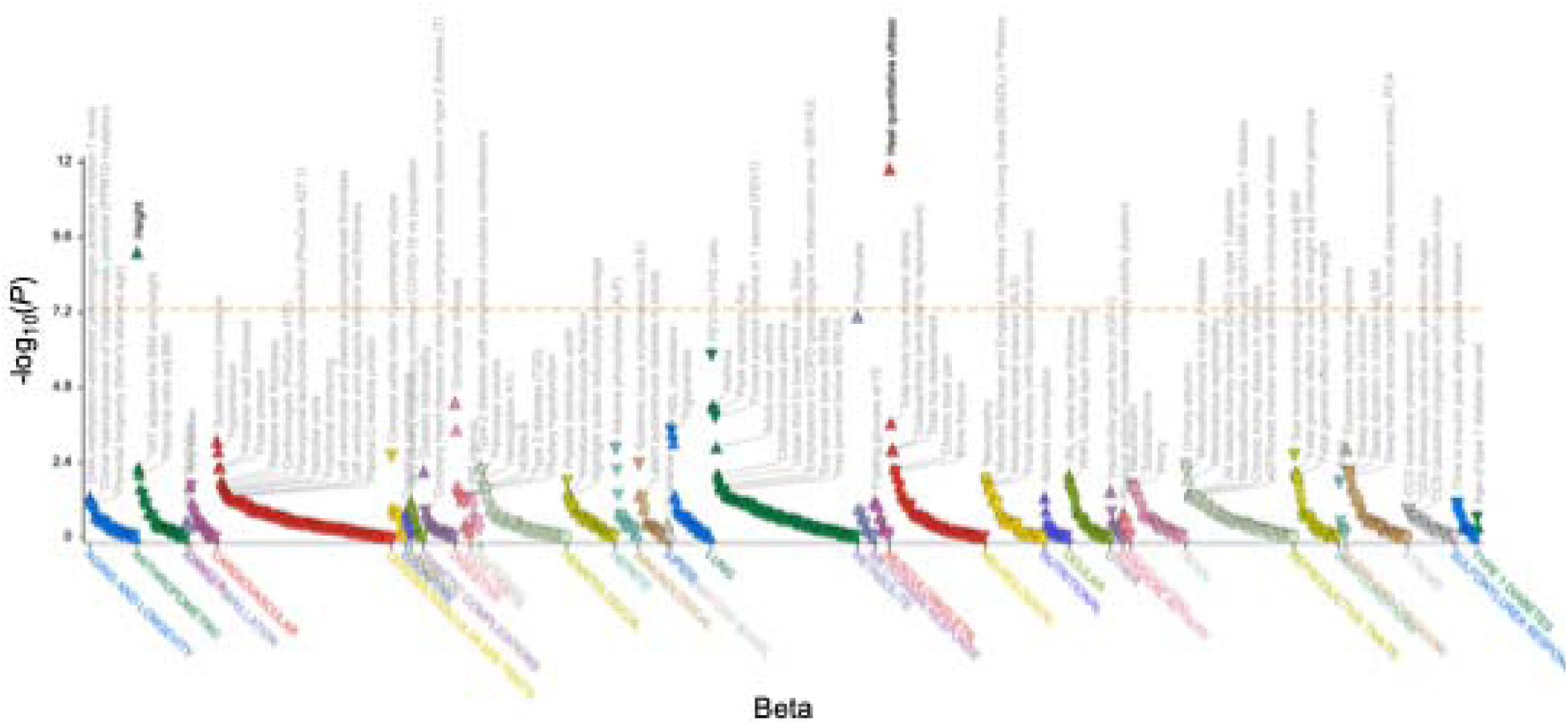
Phenome-wide association results for *TIMP2* lead *cis*-pQTL 17:76861940 C>G (rs9894212) in the deCODE cohort. The plot displays −log_10_(*P*) for the association between the deCODE lead *cis*-pQTL 17:76861940 C>G (rs9894212) and a broad range of phenotypes in the Association-to-Function (A2F) portal. Triangles denote variant-phenotype associations, grouped by phenotype category on the x*-*axis and distinguished by color. The y-axis shows −log_10_(*P*) for the association test. The horizontal orange dashed line indicates the phenome-wide significance threshold used in A2F; points above the line represent associations significant after multiple-testing correction.

**Supplementary Figure 4.**
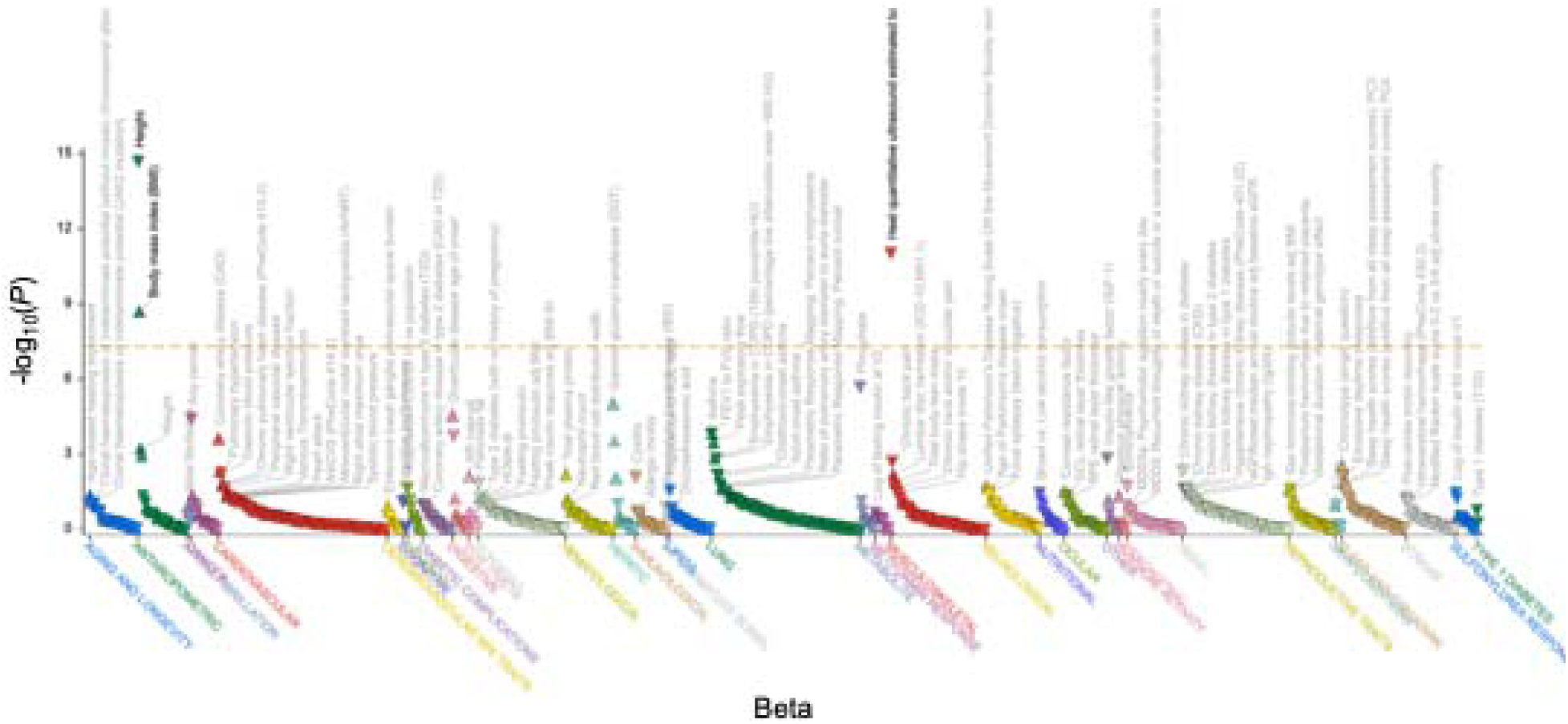
Phenome-wide association results for *TIMP2* lead *cis*-pQTL 17:76858539 T>C (rs931227) in the Fenland cohort. The plot displays −log_10_(*P*) for the association between the Fenland lead *cis*-pQTL 17:76858539 T>C (rs931227) and a broad range of phenotypes in the Association-to-Function (A2F) portal. Triangles denote variant-phenotype associations, grouped by phenotype category on the x-axis and distinguished by color. The y-axis shows −log_10_(*P*) for the association test. The horizontal orange dashed line indicates the phenome-wide significance threshold used in A2F; points above the line represent associations significant after multiple-testing correction.

**Supplementary Figure 5.**
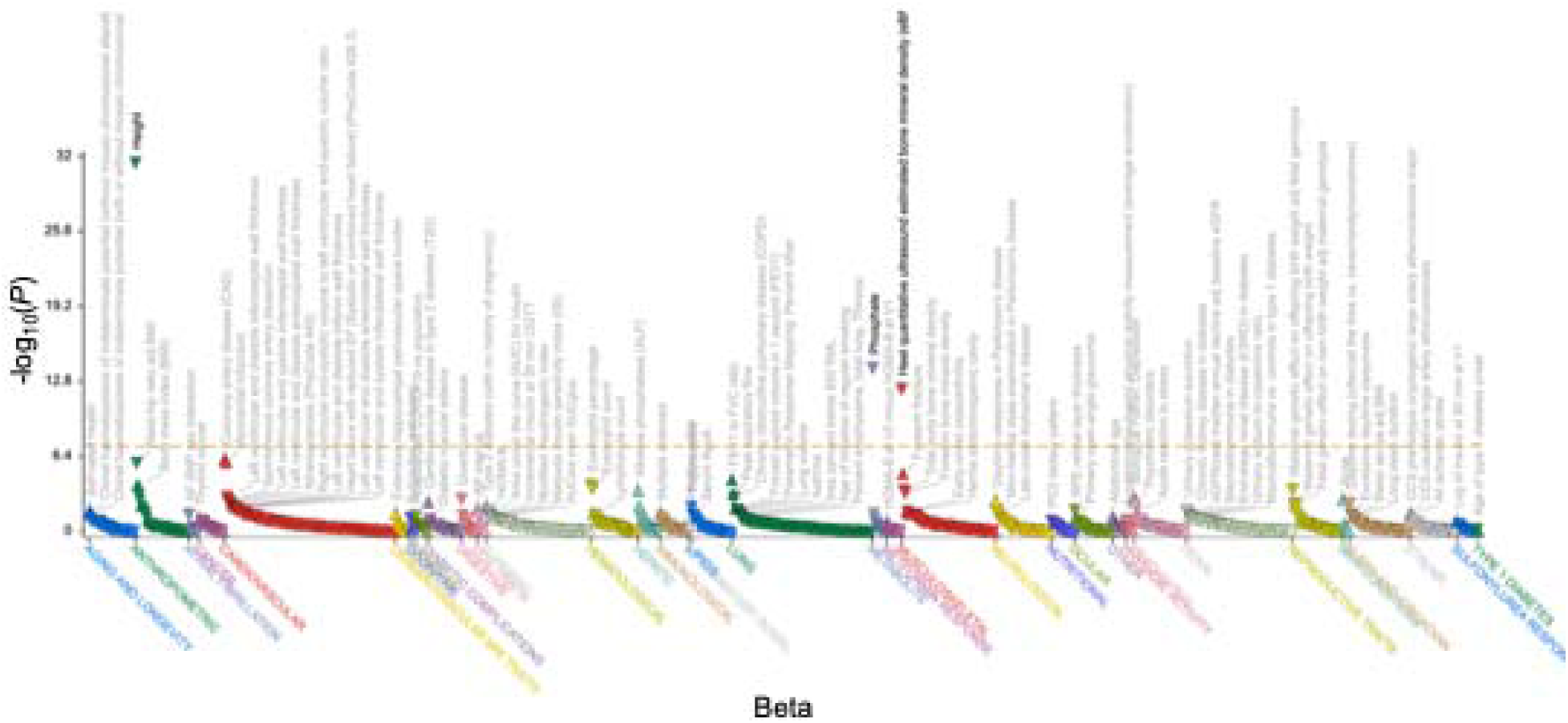
Phenome-wide association results for *TIMP2* lead *cis*-pQTL 17:76913843 T>C (rs8066695) in the UKB-PPP cohort. The plot displays −log_10_(*P*) for the association between the UKB-PPP lead *cis*-pQTL 17:76913843 T>C (rs8066695) and a broad range of phenotypes in the Association-to-Function (A2F) portal. Triangles denote variant-phenotype associations, grouped by phenotype categories on the x-axis and distinguished by color. The y-axis shows −log_10_(*P*) for the association test. The horizontal orange dashed line indicates the phenome-wide significance threshold used in A2F; points above the line represents associations significant after multiple-testing correction.

